# Oculomotor analysis to assess brain health: preliminary findings from a longitudinal study of multiple sclerosis using novel tablet-based eye-tracking software

**DOI:** 10.1101/2023.07.02.23292151

**Authors:** Étienne de Villers-Sidani, Patrice Voss, Natacha Bastien, J. Miguel Cisneros-Franco, Shamiza Hussein, Nancy E. Mayo, Nils A. Koch, François Blanchette, Daniel Guitton, Paul S. Giacomini

**Affiliations:** Innodem Neurosciences, Montreal, QC, Canada; Montreal Neurological Institute, McGill University, Montreal, QC, Canada; Novartis Pharmaceuticals Canada Inc., Montreal, QC, Canada; School of Physical and Occupational Therapy, Faculty of Medicine, McGill University, Montreal, QC, Canada; Integrated Program in Neuroscience, McGill University, Montreal, QC, Canada

## Abstract

A growing body of evidence supports the link between eye movement anomalies and brain health. Indeed, the oculomotor system is composed of a diverse network of cortical and subcortical structures and circuits that are susceptible to a variety of degenerative processes. Here we show preliminary findings from the baseline measurements of an ongoing longitudinal cohort study in MS participants, designed to determine if disease and cognitive status can be estimated and tracked with high accuracy based on eye movement parameters alone. Using a novel gaze-tracking technology that can reliably and accurately track eye movements with good precision without the need for infrared cameras, using only an iPad Pro embedded camera, we show that several eye movement parameters significantly correlated with clinical outcome measures of interest. Eye movement parameters were extracted from fixation, pro-saccade anti-saccade, and smooth pursuit visual tasks, whereas the clinical outcome measures were the scores of several disease assessment tools and standard cognitive tests such as the Expanded Disability Status Scale (EDSS), Brief International Cognitive Assessment for MS (BICAMS), the Multiple Sclerosis Functional Composite (MSFC) and the Symbol Digit Modalities Test (SDMT). Furthermore, multiple regression analyses show that a small set of oculomotor parameters can explain up to 74% of the variance of the clinical outcome measures. Taken together, these findings not only replicate previously known associations between eye movement parameters and clinical scores, this time using a novel mobile-based technology, but also the notion that interrogating the oculomotor system with a novel eye-tracking technology can inform us of disease severity and evolution, as well as the cognitive status of MS participants.

## Introduction

Multiple sclerosis (MS) is an inflammatory disease of the central nervous system, causing demyelination and neurodegeneration in most individuals, where progression can be gradual and subtle (Hauser et al, 2006; Trapp et al, 2008). Though historically MS has often been categorized by distinct clinical descriptors (e.g., primary progressive, relapsing-remitting), accumulating evidence suggests that the clinical course of multiple sclerosis is better considered as a continuum (Kuhlmann et al, 2023). Cognitive dysfunction is a common concomitant of MS and recent evidence suggests that the standard clinical interview and neurological examination are not sufficiently sensitive to detect cognitive impairment and more subtle motor impairments in MS (Romero et al 2015). It is in part for this reason that special task forces and expert consensus committees developed the Multiple Sclerosis Functional Composite (MSFC) and the Brief International Cognitive Assessment for MS (BICAMS), which when combined allow for more comprehensive assessments of cognitive processing speed, verbal and visuospatial memory, ambulation, and hand visuomotor coordination. Although these assessments have proven invaluable in research and clinical trial settings, their lengthy administration time makes them very difficult and impractical to use as part of the standard of care in routine clinical practice.

Researchers have consequently looked at alternative means to objectively assess the disease and cognitive status of individuals with MS, and one such recent avenue has been through the measurement of oculomotor parameters. Recent work has unequivocally shown that eye movements can reflect certain aspects of brain function and inform on the presence of neurodegeneration and cognitive impairment (Anderson TJ, MacAskill, 2013; Bueno et al, 2019; Crotty and Chwalisz, 2019; Terao et al, 2013). The link between eye movements and brain health should not be too surprising, given that eye movements are controlled by a diverse network of the brainstem and cortical structures and circuits (Goffart et al, 2018; Leigh and Zee, 2006) that are susceptible to a variety of degenerative processes (Gorges et al, 2014; Anderson TJ, MacAskill, 2013; Serra et al, 2018). Moreover, the analysis of gaze patterns and visual tasks that measure inhibition can provide insights into the integrity of various cognitive processes (Liu et al, 2021; Bueno et al, 2019; Fielding et al, 2015).

The literature for MS is particularly rich with evidence demonstrating oculomotor anomalies, such as fixation instability (Mallery et al, 2018; Nij Bijvank et al., 2019; Sheehy et al, 2020), slowed pro-saccades (Polet et al, 2020; Józefowicz-Korczyńska et al., 2008; Finke et al, 2012) with increased onset latencies (Nij Bijvank et al., 2020; Nygaard et al, 2015; Clough et al, 2018), increased anti-saccade error rates (Polet et al, 2020; Nij Bijvank et al., 2020; Clough et al, 2018; Fielding et al, 2009; Kolbe et al, 2014), and reduced slow pursuit gain (Lizak et al, 2016). Furthermore, several studies linked these anomalies to brain health via correlations between several eye movement parameters and disease or cognitive status as measured by tools such as the Expanded Disability Status Scale (EDSS), the Symbol Digit Modalities Test (SDMT) (Gajamange et al, 2019; Nygaard et al, 2015; Kolbe et al, 2014; Sheehy et al, 2020; Polet et al, 2020) and the PASAT (Fielding et al., 2009; Fielding et al., 2012; Kolbe et al., 2014).

These findings and others have led to suggestions that laboratory eye movement recordings can be extremely useful for objective and precise identification of disease status and monitoring of disease progression (Anderson TJ, MacAskill, 2013) and assist with differential diagnoses (Chalkias et al, 2021, Kassevetis et al, 2022; Armstrong, 2015). Unfortunately, the use of detailed eye movement recordings in clinical settings has been limited, due largely to the cost-prohibitive nature and limited scalability of the required specialized equipment, such as infrared eye-tracking cameras. However, Innodem Neurosciences has recently developed a patented gaze-tracking technology (Eye-Tracking Neurological Assessment: ETNA™) that can reliably and accurately track eye movements with good precision without the need for infrared cameras, using only the embedded camera of an iPad Pro. This technology allows for the precise quantification of several eye movement parameters currently only available with specialized and costly research-grade infrared eye tracking devices, such as the latency, velocity, and accuracy of saccades, and the presence of saccadic intrusions during fixation. The ETNA™ platform was recently used to replicate well-known oculomotor findings in Parkinson’s disease, in addition to showing that several eye movement parameters were significantly correlated with disease severity as assessed via the MDS-UPDRS motor subscale (de Villers-Sidani et al, 2023). These first set of findings demonstrated that this tablet-based tool has the potential to both accelerate eye movement research via affordable and scalable eye-tracking, and aid with the precise identification of disease status and monitoring of disease progression in clinical settings.

The purpose of this manuscript is to present preliminary findings from an ongoing longitudinal study following a cohort of persons with MS and healthy controls. The results presented herein were produced as the result of an interim device performance analysis using a subset of twenty oculomotor parameters (listed below in the methods section) collected from four different visual tasks (fixation, pro-saccades, antisaccade, and smooth pursuit), which were selected a priori and based on parameters for which there was evidence in the literature of either anomalous values in MS or correlations with our chosen MS-based clinical outcome indicators. The overarching goal of the study is to determine if disease and cognitive status can be estimated with high accuracy based on eye movement parameters alone, using Innodem’s patented mobile, scalable, and accessible eye-tracking technology. As a first step, we provide preliminary findings regarding the relationship between the preselected eye movement parameters and validated clinical scale scores linked to disease status and cognitive status. To assess disease and cognitive status, we used three of the most employed MS assessment tools (Expanded Disability Status Scale (EDSS) (Kurtzke, 1983), Brief International Cognitive Assessment for MS (BICAMS) (Benedict et al., 2012, Langdon et al., 2012), and Multiple Sclerosis Functional Composite (MSFC) (Rudick et al., 1997; Fischer et al., 1999) along with the Symbol Digit Modalities Test (SDMT), which together formed our four clinical outcome measures of interest.

## Methods

### Study Design and Subject Population

This cross-sectional study included 60 participants and was approved by both the Veritas and the McGill University Health Center (MUHC) research ethics boards (ClinicalTrials.gov Identifier: NCT05061953). All participants were persons with MS (49 with RRMS and 11 with SPMS; 41 female / 19 male; see Table 1 for additional demographic and clinical information) and were recruited from one of two study sites: the Montreal Neurological Clinic (MNC) and the Montreal Neurological Institute and Hospital (MNI). All data collection was performed by the clinical research units at the study sites (MNC: Genge Partners, Inc; MNI: The Clinical Research Unit at the Montreal Neurological Institute). The main inclusion criteria were adults with a confirmed diagnosis of MS with no signs of progressive increase in physical disability within the past six months and sufficient visual acuity to perform the tasks. The main exclusion criteria were the presence of comorbid neurological or psychiatric conditions, to avoid eye movement anomaly confounders, the recent start of medications known to influence ocular motor visual function (e.g., benzodiazepines) and participants who experienced an MS relapse at the time of assessment.

**Table 1.** Participant demographic data (n=60) and MS-related clinical test scores. EDSS (Expanded Disability Status Scale; BICAMS (Brief International Cognitive Assessment for MS; MSFC (Multiple Sclerosis Functional Composite); SDMT (Symbol Digit Modalities Test); RAVLT (Rey Auditory Verbal Learning Test); BVMT-R (Brief Visuospatial Memory Test-Revised); T25FW (timed 25-foot walk); 9HPT (9-hole pegboard test).

|  | **Mean (SD)** | **Median (IQR)** | **Min-Max** |
| --- | --- | --- | --- |
| **Age** | 51.0 (10.6) | 51 (44 - 58) | 26 - 74 |
| **EDSS** | 3.5 (2.0) | 2.5 (2.0 - 5.75) | 1.0 - 7.5 |
| **SDMT** | 49.7 (13.5) | 50 (41 - 59) | 22 - 80 |
| **RAVLT** | 54.2 (11.2) | 56 (48 - 62) | 20 - 72 |
| **BVMT-R** | 24.2 (6.9) | 25 (19 - 30) | 8 - 36 |
| **T25FW** | 29.4 (60.2) | 4.8 (4.2 - 9.2) | 2.8 - 180.0 |
| **9HPT** | 31.4 (31.5) | 22.1 (20.1 - 26.5) | 17.4 - 165.6 |
| **BICAMS** | 0.0 (0.9) | 0.1 (-0.5 - 0.6) | -2.3 - 1.3 |
| **MSFC** | -0.6 (2.1) | 0.2 (-0.5 - 0.5) | -6.4 - 1.3 |

### Clinical and cognitive assessments

To assess disease and cognitive status, we used three of the most commonly employed MS assessment tools: EDSS (Expanded Disability Status Scale; Kurtzke, 1983), BICAMS (Brief International Cognitive Assessment for MS; Benedict et al 2012, Langdon et al, 2012) and two subtests of the MSFC (Multiple Sclerosis Functional Composite; Rudick et al; 1997; Fisher et al, 1999). The EDSS is a validated disability score based on the neurological examination and takes approximately 20 minutes to complete and allows for the quantification of physical disability in multiple sclerosis and the monitoring of changes in the level of disability over time and was performed by a certified examiner (neurologist) to assess the level of disability.

The BICAMS consists of a test of information processing speed (SDMT), as well as tests for verbal (CVLT-II) and visual memory (BVMT-R), representing the most frequent cognitive deficits observed in MS (Benedict et al., 2006; Glanz et al., 2010; van Schependom et al., 2015). However, we substituted the CVLT-II with the RAVLT (Schmidt, 1996) due to it having a validated and normed version for French Canadians. Both tests are nearly identical and have been shown to be highly comparable for detecting learning deficits in MS (Beier et al, 2019; Beier et al, 2020). Moreover, there is precedence for replacing the CLVT-II with the RAVLT in BICAMS validation studies in non-English-speaking countries (Filser et al, 2018; Baetge et al, 2020).

The MSFC comprises quantitative functional measures of three key clinical dimensions of MS: leg function/ambulation (timed 25-foot walk test (T25-FW)), arm/hand function (9-hole pegboard test (9-HPT), and cognitive function (the PASAT test) (Cutter et al, 1999). However, here we only performed the two subtests with a motor component (T25-FW and 9-HPT) and replaced the PASAT score with the SDMT score to reduce testing time, and because SDMT was found to be a more valid and reliable measure of cognitive processing speed compared to PASAT (Sonder et al, 2014).

### Gaze-tracking experimental setup

All tests were performed using a 12.9-inch iPad Pro tablet with the ETNA^TM^ software installed, which enables simultaneous video recordings of the eyes and the presentation of visual stimuli on the screen. All participants performed four oculomotor tasks (a fixation task, a pro-saccade task, an anti-saccade task, and a smooth pursuit task, see **Figure 1**), which were preceded by a calibration step, where participants were instructed to follow a slowly moving target across the screen. Calibration and all four tasks were completed in under 10 minutes.

All tasks were performed with the tablet screen placed vertically, camera side up, and secured at eye level using a tablet pole mount. Participants were positioned approximately 45 cm from the tablet screen and were allowed to use their best-corrected visions, with glasses or lenses if necessary. Safeguards within the gaze-tracking software ensured the participant’s head was properly positioned and visible via the embedded camera, at an acceptable angle and distance from the screen.

**Figure 1.**
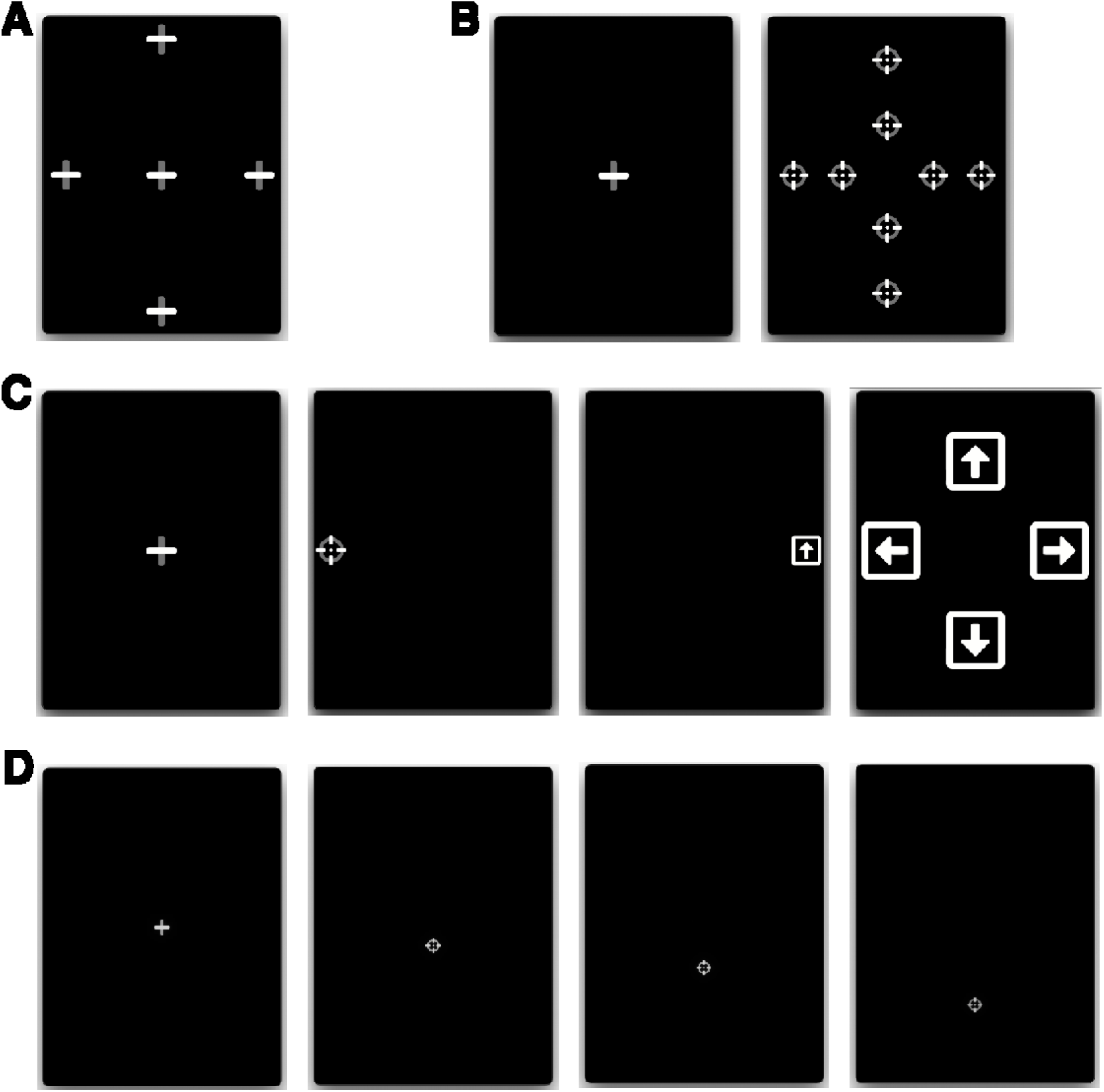
Eye-tracking tasks. (**A**) Fixation: participants fixated a stationary target for 7 seconds, at one of 5 locations. (**B**) Pro-saccades: participants initially fixated a central fixation point, which disappeared after 1.0 – 3.5 s, after which a different target appeared at one of 8 eccentric locations for 1.5 seconds. (**C**) Anti-saccades: participants initially fixated a central fixation point, which disappeared after 1.0 – 3.5 s, after which a round target appeared at 10° to the left or right from the center. Participants were instructed to move their gaze in the opposite direction to the round target, where after 1200ms they were shown a square with an arrow inside that pointed in one of 4 random directions (left, right, up, or down; shown during 400ms). The users then had to direct their gaze toward the arrow orientation corresponding to the arrow they saw in the preceding step. (**D**) Smooth pursuit: after initially fixating on a central cross, participants followed a moving target with a constant velocity of 8°/s (in this example, a downward-moving target).

#### Fixation task

Participants had to fixate a stationary target for 7 seconds, at five different locations (one central and 4 eccentric locations). The eccentric positions were located at 10 degrees of visual angle left and right from the center and 14 degrees of visual angle up and down from the center (**Figure 1A**).

#### Pro-saccade task

Participants had to initially fixate a central fixation point, which disappeared after a random period of 1.0 – 3.5 s, after which a different target reappeared at an eccentric location for 1.5 seconds either to the left or right, above or below the central fixation point. Participants were instructed to move their gaze as quickly as possible to the new target location. Both short (5° horizontal, 6° vertical) and large (10° horizontal,12° vertical) eccentric target distances were used, and each target location was sampled 3 times, for a total of 24 trials (**Figure 1B**).

#### Anti-saccade task

Participants had to initially fixate a central fixation target, which disappeared after a random period of 1.0 – 3.5 s, after which a different target reappeared at an eccentric location (10°) to the left or right from the center. Participants were instructed to move their gaze as quickly as possible in the opposite direction to the new target location. After being displayed for only 100 ms, the target disappeared, and the screen was left blank for a predetermined duration of time. Following the blank screen, a symbol appeared in the opposite location of where the initial stimulus appeared (i.e., where the participant should be looking). This symbol consisted of a white square with an arrow inside oriented in one of 4 random directions: either left, right, up, or down. The blank screen period lasted 1200 ms and the arrow symbol duration of 400ms. After each trial, a screen was displayed for 5 seconds prompting the user to answer which symbol they saw by directing their gaze toward the arrow orientation corresponding to what they believe is the correct answer (**Figure 1C**). This task was inspired by an anti-saccade task used in a previous study (Guitton et al.,1985), whereby participants could only identify the second symbol had they performed the anti-saccade task correctly (i.e., looked in the opposite direction of the initial target).

#### Smooth pursuit task

Participants here were first required to fixate a central fixation cross of variable duration (1000-2000ms). Once the fixation cross disappeared a moving target (that could either go up, down, left or right) appeared on screen for which the participants were instructed to follow with their gaze. step–ramp paradigm of smooth pursuit at constant velocity was used, whereby the initial position of the moving target was positioned offset from the central fixation point, on the opposite side of the motion direction (**Figure 1D**). For instance, in a trial of rightward smooth pursuit, the motion target would first appear to the left of the central fixation point (i.e., *the step*) and then moved in the opposite direction (rightward) at a constant velocity (i.e., *the ramp*). The trial terminated when the target reached the 10° position either left, right, above, or below center. A total of four trials were performed, one in each direction, with a constant velocity of 8°/s with a step size of 1.5°.

### Parameter extraction and analysis

Offline analysis was performed using ETNA^TM^ ‘s proprietary analysis pipeline to automatically extract the eye movement parameters reported for each task. Before parameter extraction, all gaze signals were processed and non-saccadic artifacts (e.g., blinks) were removed by the software’s analysis pipeline.

The following parameters were extracted from the ***fixation task*** gaze recordings (parameters were averaged across the five fixation trials): 1) 95% bivariate contour ellipse area (BCEA 95) of fixation – a measure of fixation stability which encompasses an ellipse that covers the 95% of fixation points that are closest to target, 2) the rate of saccadic intrusions during fixation, 3) the average amplitude of the saccadic intrusions during fixation.

The following parameters were extracted from the ***pro-saccade task*** gaze recordings (averaged across all short– and large-eccentricity targets separately): 1) average saccade latency, 2) average total time to reach the peripheral target, 3) average peak saccade velocity, 4) average saccade amplitude gain (i.e., the amplitude of the saccade relative to the eccentricity of the target; a measure of saccade accuracy).

The following parameters were extracted from the ***anti-saccade task*** gaze recordings: 1) direction error rate, 2) direction correction rate (proportion of trials where participants directed their gaze in the correct direction following an initial saccade in the wrong direction),3) correct direction saccade onset latency, 4) incorrect direction saccade onset latency, and 5) time-to-correct latency (time elapsed between a first incorrect saccade and a second corrective saccade in the right direction).

The following parameters were extracted from the ***smooth pursuit task*** gaze recordings: 1) pursuit velocity gain, 2) pursuit lag – average distance separating the target from the pursuit gaze point, 3) proportion of time spent in pursuit – the ratio of time spent in pursuit to the total time elapsed between eye movement onset and offset, relative to the time spent performing catch-up saccades and 4) number of catch-up saccades.

For all correlations between eye movement parameters and the clinical outcome measures of interest (EDSS, SDMT, BICAMS, and MSFC) the Spearman’s ñ correlation coefficient was calculated. Data analyses were performed using SAS statistical software suite. Corrected p-values to adjust for the false discovery rate were computed using the Benjamini-Hochberg procedure evaluated at an alpha level of 0.05 (Benjamini & Hochberg, 1995). Data visualization was performed using R 4.2.1 in RStudio (build 554), packages dplyr, tidyverse, ggplot2, ggpubr, and rstatix. The scores on SDMT, RAVLT, and BVMT-R were converted to z-scores and their average was calculated to obtain the composite BICAMS score. Similarly, the scores on SDMT, T25FW, and 9HPT were converted to z-scores and their average was calculated to obtain the composite MSFC score (the reciprocal of the 9HPT score was used here and the T25FW score was multiplied by –1 so that higher scores on each test corresponded to better scores).

### Comparisons of high vs low EDSS

For each oculomotor parameter, data were z-scored and then split based on high vs. low EDSS scores (EDSS ≤ 4 and EDSS ≥ 4.5) for visualization in **Figure 3** and exploratory analyses — Mann– Whitney U tests were used for comparing each ocular parameter value between the high and low EDSS groups.

### Multiple regression analyses

Multiple regression analysis was used to examine the relationship between the features and each clinical score (EDSS, SDMT, MSFC and BICAMS). An exhaustive feature selection procedure was used to select the parameters that most contributed to the final model. This procedure involved sampling all possible combinations of oculomotor parameters (set sizes from 1 to all parameters) and subsequent linear model fitting. For each set of oculomotor parameters, only patient samples with the full set of parameter values (i.e., not containing any missing values within the specific set of parameters, which for the most part were structurally missing, such as the *antisaccade correct direction latency parameter* in cases where participants did perform any correct direction anti-saccade trials) were used (n = 29 for EDSS and SDMT and n = 30 for MSFC and BICAMS) for fitting of the linear model. For each model, standardized regression coefficients were computed by multiplying regression coefficients by the standard deviation of the predictor variable divided by the standard deviation of the dependent variable. The normalized absolute values of the standardized regression coefficients are used as a measure of oculomotor parameter contribution to the model. The coefficient of determination (R2) was used to assess multiple regression performance (both adjusted and non-adjusted values) and was used as the feature selection metric. Multiple regression analyses and radar plots were conducted using scikit-learn 1.2.2 and matplotlib 3.7.1 in Python 3.9.6.

## Results

Spearman correlations between the extracted eye movement parameters and clinical outcome measures (SDMT, BICAMS, MSFC, and EDSS) are shown in **Table 2** – see **Figure 2** for a graphical representation of select representative correlations highlighting the parameter for each task that most strongly correlated with all clinical outcome measures. After correction for multiple comparisons, *nine* eye-movement parameters were significantly correlated with the SDMT, *five* with BICAMS, *ten* with the MSFC, and *nine* with the EDSS. A greater percentage of pro-saccade (56%) and anti-saccade (44%) parameters were found to significantly correlate with the MS-related clinical scale scores than did the fixation (25%) and smooth pursuit (6%) parameters.

**Figure 2.**
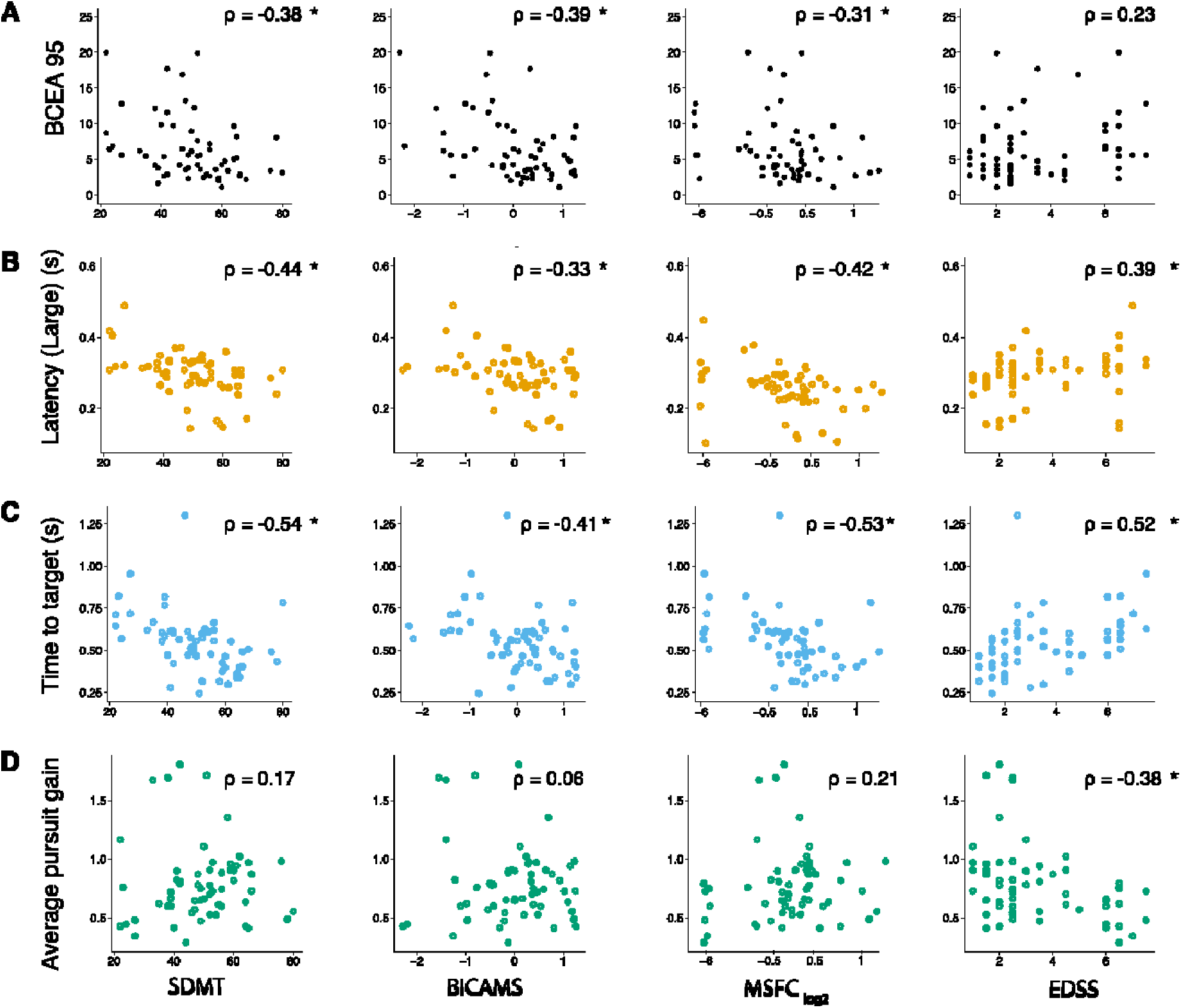
Spearman correlations between select eye-tracking parameters and functional scores. (A) Fixation: BCEA95, (B) Pro-Saccades: large amplitude saccade latency, (C) Anti-Saccades: time to target, (D) Smooth pursuit: average pursuit gain. All Spearman’s rho correlation values were calculated using the raw data. For visualization purposes only, the MSFC x-axes were rescaled [0.1–0.9] and log2-transformed. * p < 0.05 (corrected for multiple comparisons).

**Table 2.** Correlations between eye-movement parameters from the four eye-tracking task categories and the MS-related clinical outcome measures of interest. S, short amplitude pro-saccade; L, large amplitude pro-saccade. EDSS (Expanded Disability Status Scale; BICAMS (Brief International Cognitive Assessment for MS; MSFC (Multiple Sclerosis Functional Composite); SDMT (Symbol Digit Modalities Test). Corrected p-values correspond to the Benjamini-Hochberg adjusted p-values. * p < 0.05, ** p < 0.01, *** p < 0.001

We furthermore performed an additional *post-hoc* exploratory analysis to determine which oculomotor parameters best distinguished high EDSS from low EDSS participants (results are shown in **Figure 3** in the form of a radar plot). After z-scoring the parameter values relative to the entire group, high and low EDSS subgroups were compared and significant differences were found for almost all the same oculomotor parameters for which there was a significant correlation with EDSS, except for the two pro-saccade latency parameters and the antisaccade time-to-correct parameter, primarily those for which the monotonic relationship with the EDSS was the weakest amongst those that were significant.

**Figure 3.**
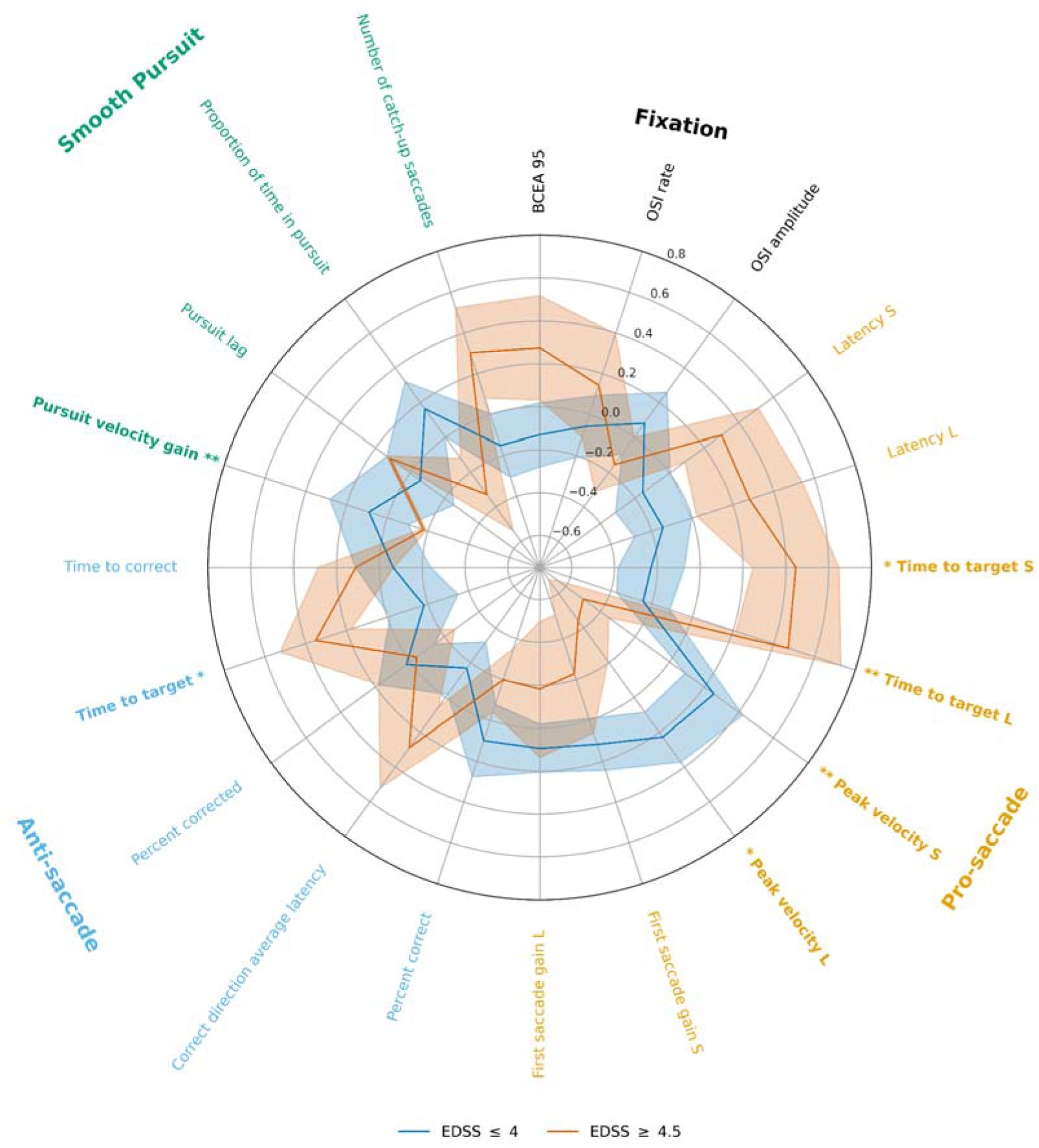
Radar plot illustrating the z-score values for low– and high-EDSS subgroups for each oculomotor parameter. Significant subgroup differences are highlighted with bold labels. S, short amplitude pro-saccade; L, large amplitude pro-saccade. * p < 0.05, ** p < 0.01.

Finally, to further assess the potential of using oculomotor parameters to estimate clinical outcome indicators, we performed multiple regression analyses for each clinical outcome measure using all the oculomotor parameters as predictors. Results are presented in **Figure 4A-D** and **Supplementary Table 1** and show that all models explain upwards of 53% of the variance of the clinical outcome measures, and up to 74% for EDSS specifically. The bottom panel of **Figure 4E** further illustrates the relative contribution of each oculomotor parameter to each model predictor. Whereas several parameters contribute to all four models (e.g., OSI rate and antisaccade time-to-target), several others are specific to one or two models (e.g., anti-saccade percent correct and smooth pursuit velocity gain).

**Figure 4.**
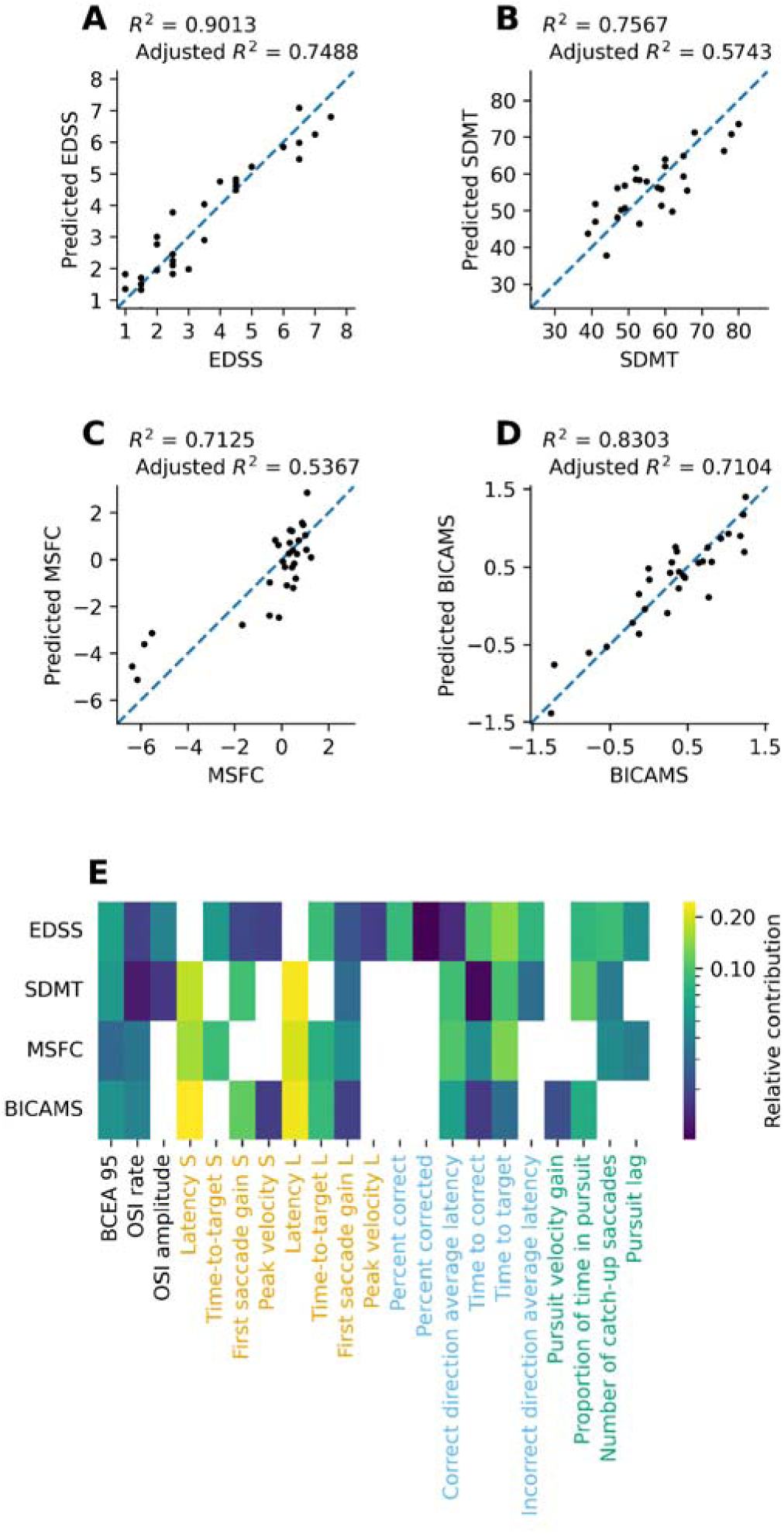
A-D) Scatterplots of the relationship between the study participants’ clinical scores and the corresponding predicted value obtained by multiple regression analysis using the oculomotor parameters as predictors. E) Heatmap visualization of the relative contribution (normalized absolute value of standardized regression coefficients) of each oculomotor parameter to each multiple regression predictor. Dark squares indicate lesser contributions to the model whereas lighter/yellow squares indicate greater contributions. Absent squares indicate that the parameter was not used in the final model. S, short amplitude pro-saccade; L, large amplitude pro-saccade.

**Supplementary Table 1.** Multiple regression tables for EDSS, SDMT, MSFC, and BICAMS. Regression coefficients (B), standard error, standardized regression coefficients (ß), t-statistic and associated p-value for each oculomotor parameter. Non-adjusted and adjusted coefficient of determination (R2 and Adj. R2) as well as the F-statistic and associated p-value for each multiple regression model.

## Discussion

The purpose of this manuscript is to present preliminary findings from an ongoing longitudinal clinical trial following a cohort of persons with MS and to demonstrate whether this novel and scalable tablet-based eye-tracking technology can provide estimates of all these clinical scales used to assess physical and cognitive MS disability. These study-specific aims were developed with the long-term goal of the trial in mind, that is to determine if disease and cognitive status can be estimated with high accuracy using a mobile, scalable, and accessible eye-tracking technology. Although we not only show that individual oculomotor findings are very much in line with those previously reported in the scientific literature on MS, which will be further discussed below, the findings from the multiple regression analyses strongly suggest that with a greater sample size and the development of ML-based tools, we may be able to accurately estimate disease severity in MS patients based on eye movement analysis alone across the full EDSS range. Indeed, the linear models explained between 53% (MSFC) and 74% (EDSS) of the variance of the clinical outcome measures using only the most predictive subset of parameters. Taken together, these findings suggest that combining different oculomotor parameters together can be very informative of an MS patient’s current functional and cognitive state, as measured by the various clinical scales in the present study. Moving forward, we believe these could serve as the building blocks required to enable more sophisticated machine learning models to estimate with a high degree of accuracy the clinical state of a patient.

As highlighted above, our oculomotor-clinical scale score correlations are consistent with those previously reported in the literature, where available. To our knowledge, no studies to date have reported correlations between eye movement parameters and the composite BICAMS or MFSC scores (with one exception, see below), but rather report correlations with one of several of their subtests, such as the T25-FW, 9-HPT, PASAT, (substituted with the SDMT here), BVTM-R, SDMT, and the CVLT (substituted with the RAVLT) here. Although at first glance the BICAMS may seem less related to oculomotor measures given the fewer number of significant correlations, it should be noted that five correlations had corrected p-values between 0.05 and 0.08 (all coefficients ≥ 0.27), suggesting that the BICAMS does indeed correlate with similar oculomotor measures, just to a lesser extent than the other examined clinical outcome measures. Finally, several parameters were found to correlate with all outcome measures – this is perhaps not so surprising given that the SDMT score was used to calculate both the BICAMS and MSFC composite scores and the known correlation between SDMT and EDSS (also significant here; Spearman rho = – 0.437, p< 0.001).

To our knowledge only one other study examined the relationship between clinical scale scores and measures of fixational stability, Sheehy et al., (2020) showed that the average number of microsaccades duration fixation increased as a function of the EDSS (r = 0.35); however, both Polet et al., (2020) and Nij Bijvank et al., (2019) indirectly supported the existence of the relationship by splitting their sample in two and showing that individuals with high EDSS scores made a greater number of saccadic intrusions than those with lower scores. Sheehy et al., (2020) furthermore showed that the average number of microsaccades was negatively correlated with the SDMT (r = – 0.35) and positively correlated with the 9HPT non-dominant hand (r = 0.39) (r = 0.35) (the authors did not find correlations with T25FW, PASAT, or 9HPT dominant hand). Although the technology used here does not capture microsaccades, our measure of fixation stability (BCEA95) showed similar correlation coefficients with SDMT, BICAMS (which includes the SDMT) and MSFC (which includes both the SDMT and 9HPT) (see **Table 2**)

With regards to the relationship between pro-saccade parameters and MS-related clinical outcome measures, Polet et al (2020) showed that individuals with high EDSS had reduced pro-saccade peak velocity compared to those with low EDSS, whereas Nij Bijvank et al., (2020) showed that pro-saccade latency was increased in individuals with high EDSS compared to those with low EDSS. Both of these findings are aligned with the negative significant correlation between peak velocities and EDSS and the positive significant correlation between latencies and EDSS observed in the present study. Zangemeister et al., (2020) showed that 9-HPT negatively correlated with peak velocity (r = – 0.321), which is consistent with the present MSFC findings (note that here the correlation sign is reversed as the higher MSFC score is better, unlike the 9-HPT). Nygaard et al., (2015) showed a negative correlation between saccade latency and SDMT r = –0.32), which is again consistent with the present findings. Finally, although Finke et al (2012) surprisingly found no correlation between the MFSC composite score and either of the peak velocity, amplitude and latency pro-saccade parameters, they did find significant correlations between the FSS score (a subscore of the EDSS) with both the latency (r = 0.385) and peak velocity (r = –0.468) parameters. While the later findings are in line with the present ones, the discrepancy regarding the MSFC correlations could be explained by the fact that we replaced the PASAT with the SDMT in our MSFC composite. Indeed, the PASAT and SDMT are moderately correlated with one another and are hypothesized to depend on different, although partially overlapping, cognitive processes (Berrigan et al., 2022).

Regarding anti-saccade parameters, Nij Bijvank et al., (2020) showed that anti-saccade latency was increased in individuals with higher EDSS scores compared to those with lower scores, whereas Gajamange et al., (2019) showed a moderate correlation (r = –0.37) between anti-saccade latency and the SDMT (though the correlation did not reach statistical significance due to a small sample size). Here we found that the correct direction anti-saccade latency significantly correlated with not only the SDMT but also the BICAMS and MSFC, whereas the correlation with EDSS just failed to meet the criteria for significance after correction for multiple comparisons. Although the anti-saccade error rate has been shown to significantly correlate with the SDMT ((r = – 0.48) Gajamange et al., (2019); (r = –0.66) Kolbe et al., (2014)), here the correlation between the error rate (or percent correct rate) and both BICAMS and SDMT failed to meet the criteria for statistical significance following correction for multiple comparisons. Although no studies investigated direction correlations between anti-saccade error rate and the EDSS, Polet et al (2020) showed that individuals with a high EDSS had increased antisaccade error rates than those with low EDSS. Here, however, there was no significant correlation between the percent correct rate and the EDSS. Finally, the two strongest links we found across all task parameters with the MS-related outcome measures have not been, to our knowledge, previously demonstrated. There were significant correlations between all outcome measures and two anti-saccade parameters: time-to-correct and time-to-target. The latter is a measure of the time elapsed between the onset of target appearance and the end of the final saccade, whereas the former is the time elapsed between an initial incorrect saccade and the initiation of a secondary corrective saccade in the opposite direction.

Regarding smooth pursuit, Lizak et al., (2016) showed that there was a strong correlation between the onset latency to the slowest velocity and the EDSS (r=0.31) — a parameter not measured in the present study – they did not find significant correlations between EDSS and pursuit gain, catch-up saccade count or amplitude (correlation coefficients not reported). This is in contrast to the present data where we identified a significant correlation between the EDSS and the pursuit gain. Whereas we found no other significant correlations between any other parameter and either of the clinical scores, Rempe et al (2021) showed both that T25-FW and EDSS significantly correlated with pursuit onset latency (r=0.478; r= 0.564), pursuit gain (r= –0.519; r= –0.484), and proportion time in pursuit (r = –0.527; r = –0.673) – no correlations were found for the average amplitude of the catch-up saccades. It is currently unclear as to why we did not find as many or as strong correlations in the present study. One possible explanation relates to the velocity of the pursuit stimulus used — while both studies use a step-ramp pursuit paradigm, the velocities reported by Rempe et al., (2021) were in the 16-24 deg/s range, whereas here we report findings for a pursuit velocity of 8 deg/s. Fortunately, we should be able to address this issue in the future as we have collected data with higher pursuit target velocities in this study, but parameters measured at these velocities were not selected as part of the a priori parameters to be examined in this preliminary study report.

The present study is not without limitations. Among them are the small sample size and the use of a small number of oculomotor parameters to investigate relationships with, and derive estimates of, clinical scale scores. Upon completion of the study, the sample size will have more than doubled and the number of investigated oculomotor parameters will have significantly increased as well. Another limitation relates to the long-term goal of being able to accurately estimate clinical scale scores in a given individual based on oculomotor parameters. Although here we show very promising results via the multiple linear regression analyses that suggest that it may be possible to do so, such analyses produce inference models, which do not guarantee strong predictive abilities. Indeed, to be able to confidently claim that we can estimate disease status or severity in a single individual, predictive models need to be validated with an independent dataset. Another potential limitation is the inclusion of the SDMT score in the MSFC composite calculation. Although there is a precedent for doing so (e.g., Drake et al, 2010), it will likely limit direct comparability with both past and future studies using the traditional MSFC score composite. We nonetheless felt the PASAT had important limitations in deciding not to include it in the current trial, including a lower test-retest reliability than the SDMT and important ceiling effects (Sonder et al, 2014), and limited use outside of clinical trials due to both being a lengthy assessment time and requiring specialized audio equipment not routinely available in clinics (Cossburn et al., 2012).

To conclude, this cross-sectional study shows promising correlations between individual oculomotor parameters and validated clinical assessment scale scores, similar to previously published studies using research-grade eye trackers. When completed, this trial will hopefully demonstrate the reliability of mobile oculomotor assessments for the monitoring of MS progression as a non-invasive, accessible, scalable and sensitive novel digital biomarker of disease progression – both for cognitive and physical disability. Indeed, the results of the multiple regression analyses show that a large portion of the clinical outcome measure variance can be explained using only a subset of oculomotor parameters. The next steps in this project will focus on integrating machine learning models comprising combinations of multiple oculomotor parameters to optimize the reliability and accuracy of the clinical scale estimates in MS patients.

## Data Availability

All data produced in the present study are available upon reasonable request to the authors.

## Notes

### Competing Interest Statement

E.d.V.-S. is a co-founder of Innodem Neurosciences, which developed the Eye-Tracking Neurological Assessment (ETNA) technology used in this study. P.V. has ownership options in Innodem Neurosciences. J.M.C.-F. is a part-time employee of Innodem Neurosciences and N.A.K. is a research intern at Innodem Neurosciences. D.G. is an unpaid consultant of Innodem Neurosciences. N.B., S.H. and F.B. are employees of Novartis Pharmaceuticals Canada Inc. P.G. is a paid medical and scientific advisor with Innodem Neurosciences and has stock options in Innodem Neurosciences. The authors declare that the research was conducted in the absence of any other commercial or financial relationships that could be construed as a potential conflict of interest.

### Clinical Protocols

https://classic.clinicaltrials.gov/ct2/show/NCT05061953

### Funding Statement

This project was supported by Natural Science and Engineering Research Council of Canada, grant RGPIN-2019-04761.

### Author Declarations

Ethics committee of McGill University Health Cente gave ethical approval for this work. Veritas IRB gave ethical approval for this work.

